# Host genetic variants near the *PAX5* gene locus associate with susceptibility to invasive group A streptococcal disease

**DOI:** 10.1101/19003087

**Authors:** Tom Parks, Kathryn Auckland, Theresa L. Lamagni, Alexander Mentzer, Katherine Elliott, Rebecca Guy, Doreen Cartledge, Lenka Strakova, Daniel O’Connor, Andrew J Pollard, Stephen J. Chapman, Matthew Thomas, Malcolm Brodlie, Julien Colot, Eric D’Ortenzio, Noémie Baroux, Mariana Mirabel, James J. Gilchrist, J. Anthony G. Scott, Thomas N. Williams, Julian Knight, Andrew C. Steer, Adrian V. S. Hill, Shiranee Sriskandan

## Abstract

We undertook a genome-wide association study of susceptibility to invasive group A streptococcal (GAS) disease combining data from distinct clinical manifestations and ancestral populations. Amongst other signals, we identified a susceptibility locus located 18kb from *PAX5*, an essential B-cell gene, which conferred a nearly two-fold increased risk of disease (rs1176842, odds ratio 1.8, 95% confidence intervals 1.5-2.3, P=3.2×10^−7^). While further studies are needed, this locus could plausibly explain some inter-individual differences in antibody-mediated immunity to GAS, perhaps providing insight into the effects of intravenous immunoglobulin in streptococcal toxic shock.

## Main

Invasive group A streptococcal (GAS) disease is defined by the isolation of *Streptococcus pyogenes* from a normally sterile site. While various manifestations can occur, infections of the skin and soft tissues are most frequent. Relative to other infections, invasive GAS is associated with a high case fatality rate[1], with an estimated 163,000 deaths attributable to the condition each year worldwide.[2]

Over time the majority of the population experience some degree of exposure to GAS yet invasive infections are rare.[3] Moreover, while the majority of invasive infection occur at the extremes of age, it remains unclear why a minority of otherwise healthy children and young adults develop devastating invasive infections.[4] While several pathogen and environmental factors are implicated, a role for host genetic variation is certainly plausible given the heritability of other GAS diseases, including rheumatic fever[5] and recurrent pharyngitis.[6] Indeed, host genetic variation in the human leukocyte antigen (HLA) locus has previously been linked to susceptibility to invasive GAS disease,[7,8] perhaps reflecting preferential binding of certain alleles to GAS superantigens.[8-10] Additionally, a number of reports have now emerged linking host genetic variation elsewhere in the genome to susceptibility to invasive bacterial disease more broadly.[11-13] Accordingly, we undertook a genome-wide association study (GWAS) of susceptibility to invasive GAS disease, combining the results of four case-control studies covering distinct clinical manifestations and ancestral populations.

This study used genotyping data from several projects, brought together to form four case-control analyses (Suppl. Figure 1). Ethical approval was granted to each project by the relevant institutional authorities.

**Figure 1.**
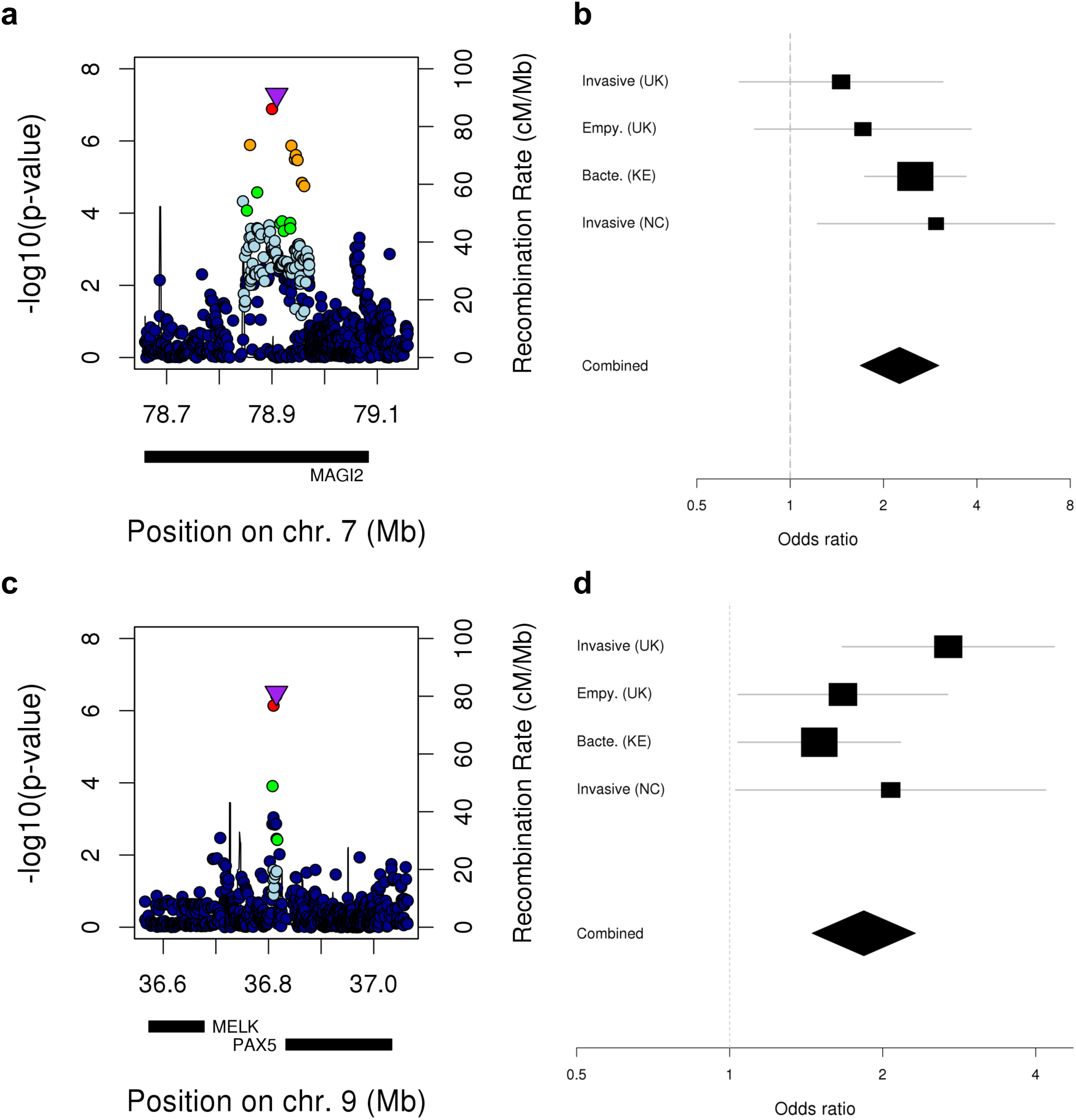
Association of the *MAGI2* and *PAX5* loci with invasive GAS disease. **(a)** Regional association for the *MAGI2* locus with **(b)** a forest plot for rs35089407, the lead variant. **(c)** Regional association plot for the *PAX5* locus with **(d)** a forest plot for rs1176844. In the regional association plots, genomic position is plotted against the negative common logarithm of the p-value from the fixed-effects meta-analysis for all variants within 250kb of the variant most associated with susceptibility. Variants are coloured by linkage disequilibrium with the most associated variant averaged across the entire data set (estimated r^2^: dark blue, 0– 0.2; light blue, 0.2–0.4; green, 0.4–0.6; yellow, 0.6–0.8; red, 0.8–1.0). In the forest plots, the effect size estimates for the lead variant at each locus is shown under an additive genetic model with association statistics from each analysis combined by fixed-effects meta-analysis.

In the first, cases were children and adults of European ancestry with GAS-associated necrotising fasciitis (NF) (n=34) or other manifestations of invasive GAS (n= 9) treated at hospitals across the UK. We have previously investigated the relationship between the variants in HLA locus and risk of invasive GAS disease amongst these individuals finding the *DQA1*\*01:03 associated with susceptibility.[14] Individuals aged 65 years or more or with co-morbidity were excluded. The youngest patient was 18 months of age and oldest was aged 63 years (median 35 years). In the second analysis, cases were children of European ancestry with GAS empyema thoracis (n=36) recruited from hospitals across the UK. The youngest patient was aged four months and the oldest 17 years (median 4 years). None had comorbidities or established immunodeficiency.

Both sets of cases were genotyped using the HumanCore platform (Illumina Inc., USA) or the Global Screening Array (Illumina) specifically for this analysis. Controls for both analyses were healthy adolescents of European ancestry recruited to vaccine studies based in Oxfordshire (n=1540) and had previously been genome-wide genotyped using the HumanOmniExpressExome platform (Illumina).

In the third analysis, cases were children of East African ancestry with GAS bacteraemia (n=66) recruited at a single rural hospital in Kilifi, Kenya, as part of a broader study of childhood bacteraemia in this setting. [15] The youngest patient was aged one day and the oldest nine years (median 162 days). Controls for this analysis were children recruited to a birth cohort set in the area surrounding that hospital (n=1689). Both cases and controls had been genotyped previously by the Wellcome Trust Case Control Consortium 2 using the Affymetrix 6.0 platform (Affymetrix, USA).[12,13]

In the fourth analysis, cases were children and adults without comorbidity of Oceanian ancestry with various invasive GAS phenotypes, including bacteraemia without source (n=7), soft tissue infection (n=3) and meningitis (n=2), treated at hospitals in New Caledonia (NC), a French territory in the South Pacific. The youngest patient was aged two months and the oldest patient 24 years (median three years). Controls for this analysis were healthy adult volunteers reporting Oceanian ancestry recruited in NC to a previously reported study of rheumatic heart disease (n=193).[16] Both cases and controls were genotyped using the HumanCore platform (Illumina).

Quality control was undertaken using standard approaches[17] but with an additional step to minimise confounding due to the use of multiple genotyping platforms (Suppl. Figure 1 & 2).[18] Crucially, after these steps, the resulting case-control datasets were well matched for genetic ancestry (Suppl. Fig. 3). Next, we randomly selected ten controls for each case carrying forward 161 cases and 1610 controls for analysis, equating to an effective sample size of 585 individuals. We then performed genotype imputation, run separately for the UK, Kenyan and NC datasets, before running case-control association analyses in each dataset using linear mixed models implemented in GCTA software[19] (v1.26.0) with no evidence of residual confounding based on the distribution of test statistics (Suppl. Figure 4). We then performed genome-wide meta-analysis using Metasoft software[20] (v2.0.1) for 4,234,532 variants present at minor allele frequency greater than 5% in all four datasets. Further analyses including estimation of effect sizes by transformation[21] were performed in R (v3.0). Finally, in the absence of a further dataset in which to confirm or refute our findings, we designed a three-step process to identify the most robust association signals that would be suitable for further investigation (Suppl. Fig. 5). This involved an initial screening genome-wide meta-analysis under the standard fixed-effects model followed by further analysis of putatively associated loci using the more stringent random-effects model to assess heterogeneity. Taking forward genomic regions containing at least one variant at suggestive significance in the random-effects meta-analysis (*P*<10^−5^), we performed a sensitivity analysis in which we reran the random selection of controls 100 times over before repeating the individual case-control analyses and rerunning the fixed-effects meta-analysis, prioritising those loci at which the association with susceptibility showed the greatest consistency.

In our initial genome-wide screen, we identified one genomic region that contained a variant associated with susceptibility at genome-wide significance (*P*FE<5×10^−8^) and 24 genomic regions that contained at least one variant associated at suggestive significance (*PFE*<10^−5^) (Suppl. Table 1; Suppl. Figure 5a). Thirteen of these regions also contained at least one variant at suggestive significance in the random-effects meta-analysis (Suppl. Table 2; Suppl. Figure 5b). In our sensitivity analysis, we found only five regions (nine variants) had no *P*FE values greater than 1×10^−4^ while only two regions (five variants), which we took forward for further analysis, had no PFE values greater than 5×10^−5^ (Suppl. Table 3).

The first of two regions shortlisted in the sensitivity analysis (maximum *P*FE<5×10^−5^) was on chromosome 7 in an intron of the *MAGI2* gene. The lead variant (rs35089407, *P*FE=5.3×10^−8^) was well supported by surrounding variants (Figure 1a). The effect was strongest in the NC analysis where the minor allele (thymidine) was associated with a three-fold increased risk of disease declining to a 1.5-fold increase in the UK invasive disease analysis (combined odds ratio, OR, 2.25, 95% confidence interval, CI, 1.68– 3.02; Figure 1b). The overall minor allele frequency was 14.7% (range 7.5-21.9%) while the imputation accuracy was high (information metric, 0.96–0.97).

The second region was on chromosome 9 located 18kb from the *PAX5* gene in the intron of a non-coding long RNA (termed AL450267.2). The peak compromised four neighbouring variants (*P*FE=3.2×10^−7^–7.3×10^−7^; Figure 1c) of which one was at suggestive significance in the random-effects analysis (rs1176844, *P*RE=7.6×10^−6^). The minor allele of rs1176844 (thymidine) was associated with an almost two-fold increased risk of the disease in the combined analysis (combined OR 1.9, 95% CI 1.4– 2.4; Figure 1d). Notably this variant, which had a minor allele frequency of 43.3%, had been directly genotyped in both the UK and NC datasets while the imputation accuracy in the Kenyan dataset was very high (information metric, 0.997).

We next investigated overlap between the five shortlisted variants in these two loci and known or predicted regulatory elements (Suppl. Table 3) using the RegulomeDB tool.[22] Overall evidence of regulatory function was minimal with nothing to suggest altered transcription factor binding or gene expression. However, rs1327501 within the *PAX5* signal overlapped histone modifications consistent with enhancer elements found specifically in analyses of primary hematopoietic stem cells and primary B cells from cord blood but no other cell types. Additionally, the other three *PAX5* variants overlapped an area of weak transcriptional activity in primary hematopoietic stem cells in short term culture but no other cell types (Suppl. Figure 6).

Finally, we reviewed the human leukocyte antigen (HLA) locus in light of our earlier report of an association between the *DQA1*\*01:03 allele and susceptibility in the UK invasive disease component of our dataset. [14] While we did not perform HLA imputation for this analysis, it is noteworthy that the two missense variants that define the *DQA1*\*01:03 allele (i.e. rs36219699 and rs41547417) were excluded from this analysis because their minor allele frequency was less than 5% in the Kenyan dataset. However, neither were associated with susceptibility in the UK empyema or NC analyses (*P*=0.50–0.65). Despite this, the strongest signal in the HLA locus mapped to within 1kb of *HLA-DQA1* (rs17612669, OR 1.5, 95% 1.1–2.1, *P*FE=0.005). While this signal is difficult to interpret in the context of a GWAS, it is noteworthy that it showed negligible heterogeneity despite the diverse genetic ancestry of the study population.

## DISCUSSION

This preliminary investigation of susceptibility to invasive GAS disease provides the first evidence that common host genetic variants may influence the risk of this infection across a range of populations, age groups and sub-phenotypes.

The signal located 18kb from *PAX5* is of particular interest because this gene is widely considered a key regulator of B cell maturation and fundamental to the genetic processes that lead to antibody production.[23] This is relevant because deficient antibody-mediated immunity to key GAS virulence factors has long been proposed as a key determinant of invasive GAS susceptibility in the general population. [24,25] Indeed, antibody-mediated neutralisation of virulence factors including superantigens may drive the protective effects of intravenous immunoglobulin, which, when administered in streptococcal toxic syndrome, was recently shown to be associated with a two-fold reduction in mortality at 30 days.[26] Moreover, the overlap with putative regulatory elements in haemopoietic stem cells, while far from definitive in the absence of experimental validation, raises the possibility these variants somehow impact differentiation of haemopoietic stem cells into B cell precursors, which might have downstream effects on antibody-mediated immunity.

In contrast, the *MAGI2* signal is harder to relate to invasive GAS disease. *MAGI2* plays a fundamental role in maintenance of synapses, endocytosis and postendocytic trafficking. [27] The gene is large, stretching 1.4Mb, and it is of interest that the locus has previously been to linked to infantile spasms. [28] Additionally, the gene has been linked by GWAS to traits including blood lipid and bone metabolism phenotypes[29] but overall it remains unclear how this locus could influence susceptibility to invasive GAS disease.

This study has a number of limitations, not least of which is the small sample size relative to most modern genetic association studies. Consequently, our analysis has limited power to detect all but the strongest signals while our estimates of effect size are imprecise and should be interpreted with caution. While follow-up of these preliminary findings is a vital next step, it is important to emphasise that invasive GAS is a rare disease and sporadic which renders recruitment challenging. Indeed, despite our focus on children and young adults, this report is, to our knowledge, the largest study of genetic susceptibility to invasive GAS to date.

Moreover, we deliberately set out to identify variants influencing susceptibility to GAS across a range of clinical manifestations and ancestral populations, as a consequence of which our study largely ignores the heterogeneous nature of this disease. Indeed, such heterogeneity might explain our findings in the HLA locus since it is possible the *DQA1*\*01:03 allele effects predisposition to only a subset of clinical phenotypes or is dependent on the presence of specific bacterial factors such superantigens.[14] Future larger-scale prospective studies with access to more precise clinical data coupled with information about the pathogen should help clarify this issue. Nonetheless, our use of data from three distinct ancestral populations has substantial advantages, not least of which is improved detection of underlying causal variants.[30]

Overall, this is the first time a GWAS of susceptibility to invasive GAS has been undertaken. While the small sample size necessitates caution, these results suggest common genetic variation in the *PAX5* locus, as well as potentially elsewhere in the genome, impacts susceptibility to invasive GAS disease. While follow-up of our findings in independent datasets is warranted, this analysis underscores the potential of investigating genetic susceptibility to GAS disease to bring about an improved understanding of pathogenesis.

## Data Availability

Several components of the data referred to in the manuscript are available through the European Genome-phenome Archive including: the UK Invasive GAS study data (accession number EGAS0000100342); the Kenyan Bacteraemia Study Group data (EGAS00001001756); the Pacific Islands Rheumatic Heart Disease Genetics Network data (EGAS00001001881). Other components will be submitted to the European Genome-phenome Archive over the coming months and/or for the most part be available from the authors on reasonable request.

https://www.ebi.ac.uk/ega/studies/EGAS00001003421

https://www.ebi.ac.uk/ega/studies/EGAS00001001756

https://www.ebi.ac.uk/ega/studies/EGAS00001001881

## Acknowledgments

This research was supported by grants awarded to T.P. from the Medical Research Council (G1100449), the European Society of Clinical Microbiology & Infectious Diseases (Research Grant 2013), the National Institute for Health Research (ACF-2016-20-001) and the British Heart Foundation (PG/14/26/30509). In addition, the genotyping undertaken by the Kenyan Bacteraemia Study Group was funded as part of the Wellcome Trust Case Control Consortium 2 (084716/Z/08/Z, 085475/B/08/Z and 085475/Z/08/Z). The genotyping undertaken by the Oxford Vaccine Group as part of EUCLIDS was funded through the European Union’s Seventh Framework Programme (EC-GA no. 279185). A.J.M. was supported by a Wellcome Trust Clinical PhD Fellowship (106289/Z/14/Z) while A.J.M., S.J.C., D.O. and A.J.P. were supported by the National Institute for Health Research Oxford Biomedical Research Centre, Oxford, UK. Additionally, M.B. was supported by a Medical Research Council Clinician Scientist Fellowship (MR/M008797/1), J.J.G. was supported by a Wellcome Trust Clinical PhD Fellowship (102342/Z/13/Z), J.A.G.S. was supported by a Wellcome Trust Senior Research Fellowship (098532), T.N.W. was supported by a Wellcome Trust Senior Research Fellowship (091758), J.C.K. was supported by a Wellcome Trust Investigator Award (204969/Z/16/Z) and A.V.S.H. was supported by a Wellcome Trust Senior Investigator Award (HCUZZ0) and by a European Research Council Advanced Grant (294557). S.S. was supported by the National Institute for Health Research Health Protection Research Unit in Healthcare Associated Infections and Antimicrobial Resistance, Imperial College London, London, UK.

We thank the Oxford Genomics Centre at the Wellcome Centre for Human Genetics for generating the genotyping data, subsidized by Wellcome Trust Core Awards (090532/Z/09/Z and 203141/Z/16/Z). We also thank the Oxford Biomedical Research Computing facility, a joint development between the Wellcome Centre for Human Genetics and the Big Data Institute supported by Health Data Research UK and the National Institute for Health Research Oxford Biomedical Research Centre with funding from the Wellcome Trust Core Award Grant Number (203141/Z/16/Z). We also acknowledge the support of the National Institute for Health Research Oxford Biomedical Research Centre, Oxford, the National Institute for Health Research Imperial Biomedical Research Centre, London, UK, and the National Institute for Health Research Newcastle Biomedical Research Centre, Newcastle upon Tyne, UK. None of these funders had any role in study design, data collection and analysis, decision to publish or preparation of the manuscript. Moreover, the views expressed here are those of the authors and not necessarily those of the National Health Service, the National Institute for Health Research or the Department of Health.

---

**Suppl. Figure 1.**
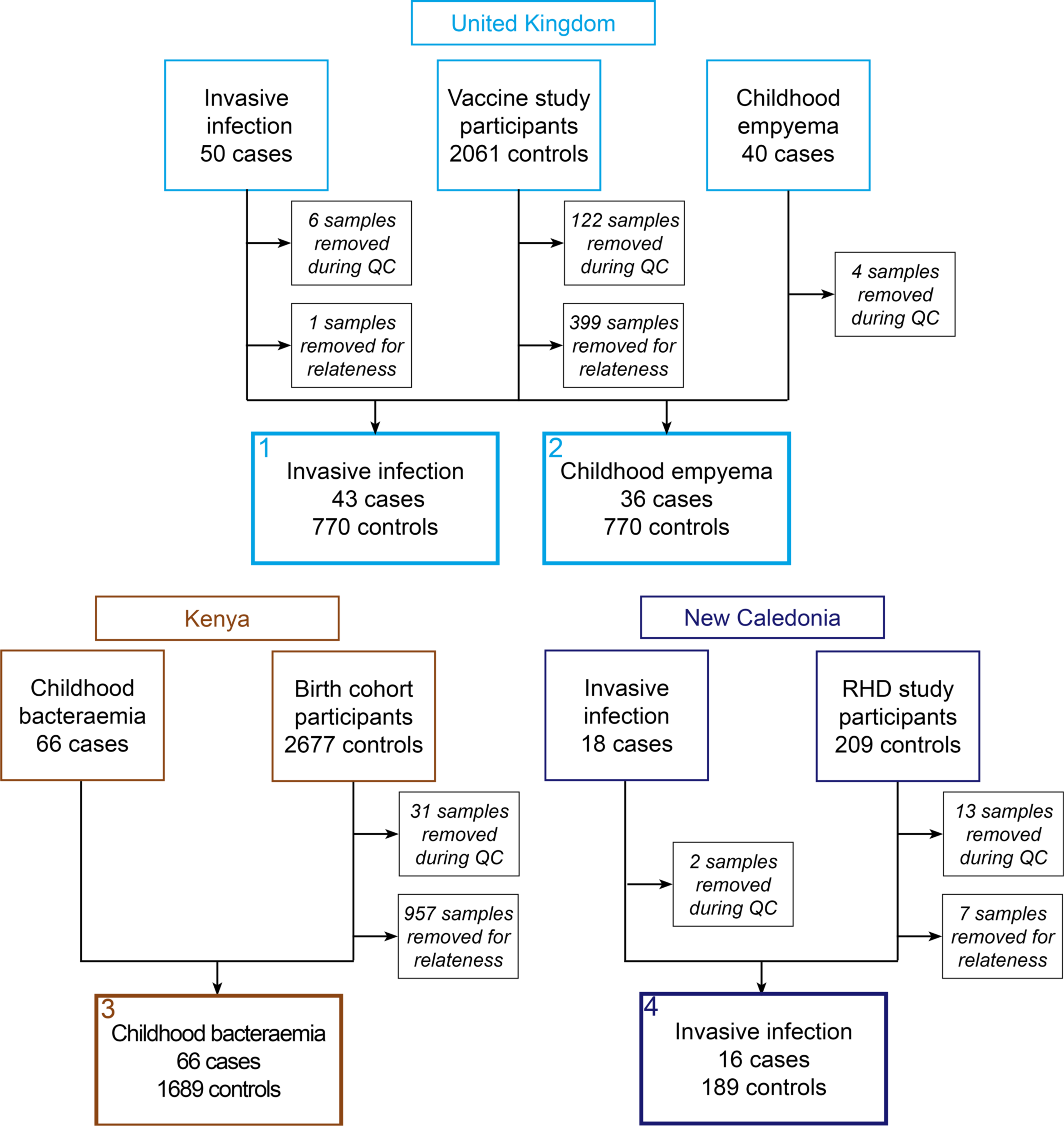
Outcome of sample quality control procedures.

**Suppl. Figure 2.**
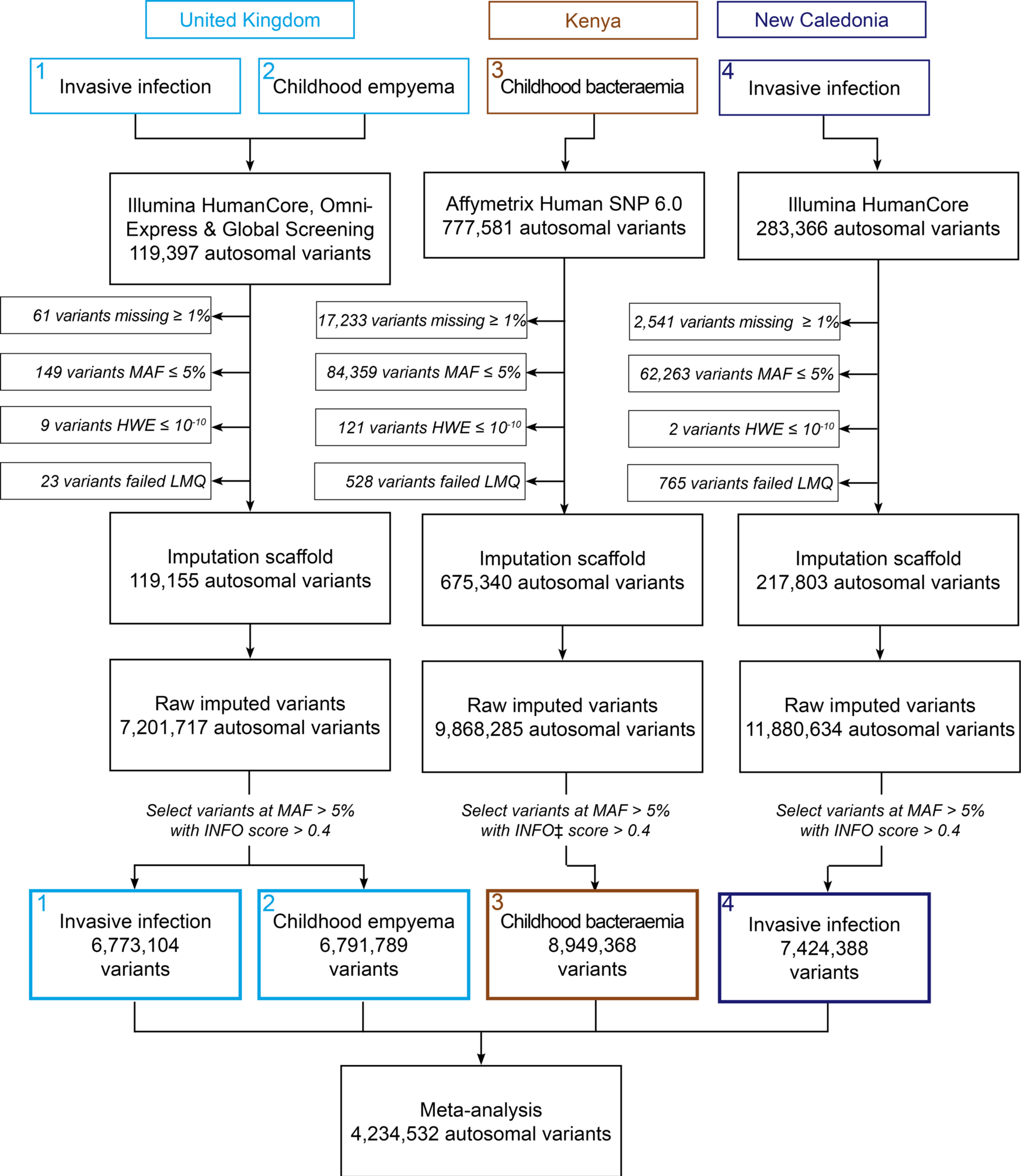
Outcome of variant quality control procedures. MAF, minor allele frequency; HWE, Hardy-Weinberg equilibrium; LMQ, two-locus linear model-based quality control test;[18] INFO, Impute2 software information metric.[31]

**Suppl. Figure 3.**
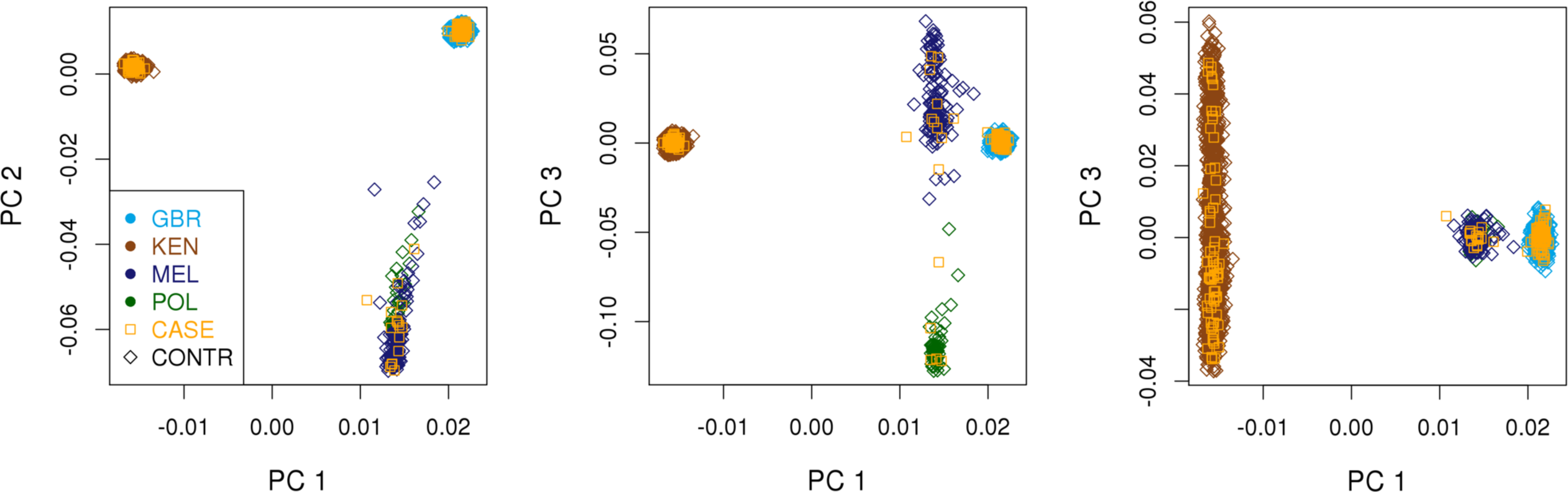
Ancestry of study population. Projection of the samples on to **(a)** the first and second, **(b)** first and third and **(c)** first and fourth principal components of genetic variation. Cases indicated by gold-coloured empty squares while controls are indicated by empty diamonds coloured by self-reported ancestry (GBR, British European; KEN, Kenyan African; MEL, Melanesian; POL, Polynesian).

**Suppl. Figure 4.**
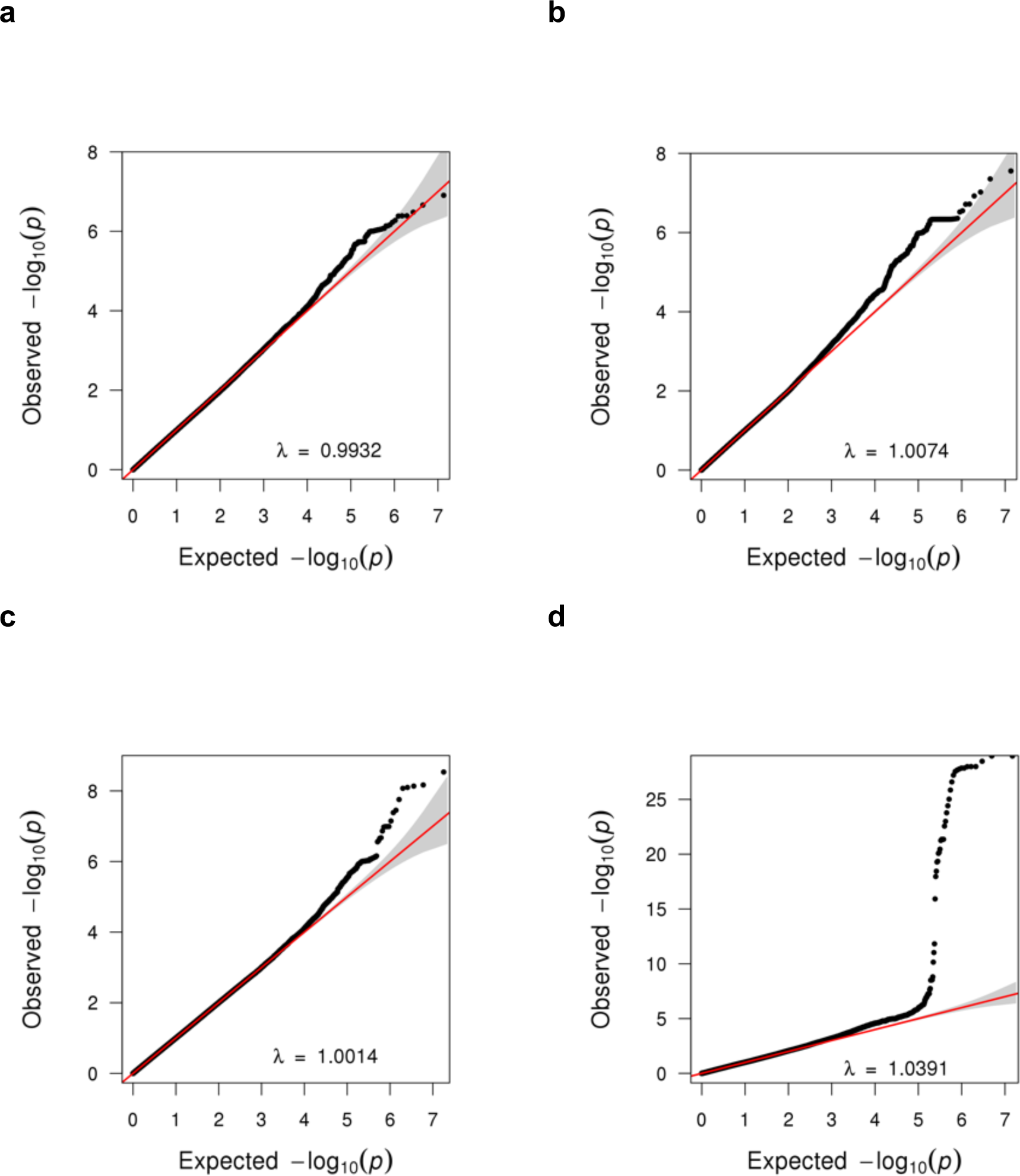
Quantile-quantile plots for individual association analyses. For each analysis, directly genotyped and imputed variants were tested for association with invasive GAS susceptibility using linear mixed models. Each point represents a single variant. An estimate of the genomic inflation factor (λ) is shown. Plots are shown for: **(a)** Invasive infection, UK, **(b)** Childhood empyema, UK, **(c)** Childhood bacteraemia, Kenya, and **(d)** Invasive infection, New Caledonia.

**Suppl. Fig 5.**
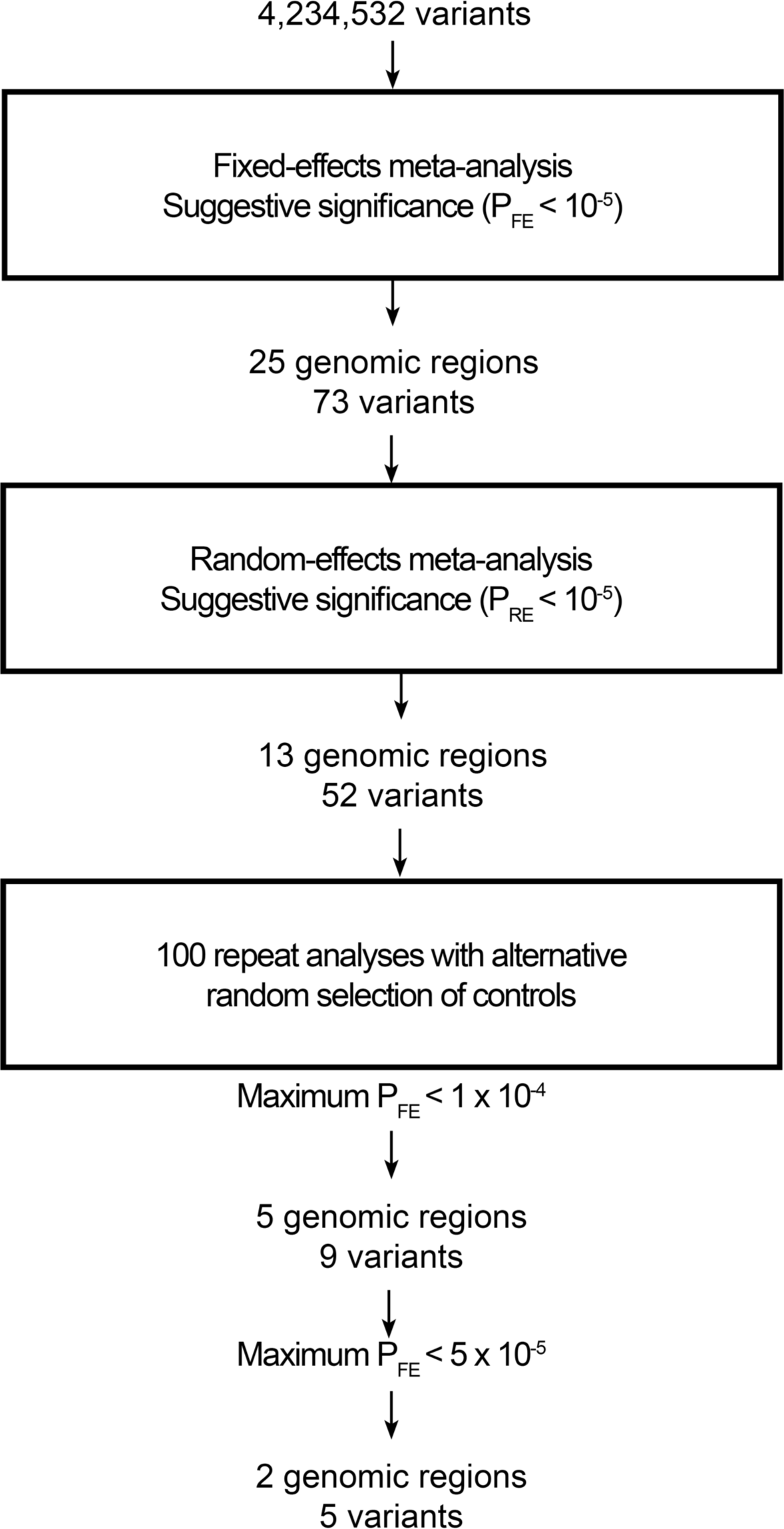
Three-step prioritisation of variants for further investigation.

**Suppl. Figure 6.**
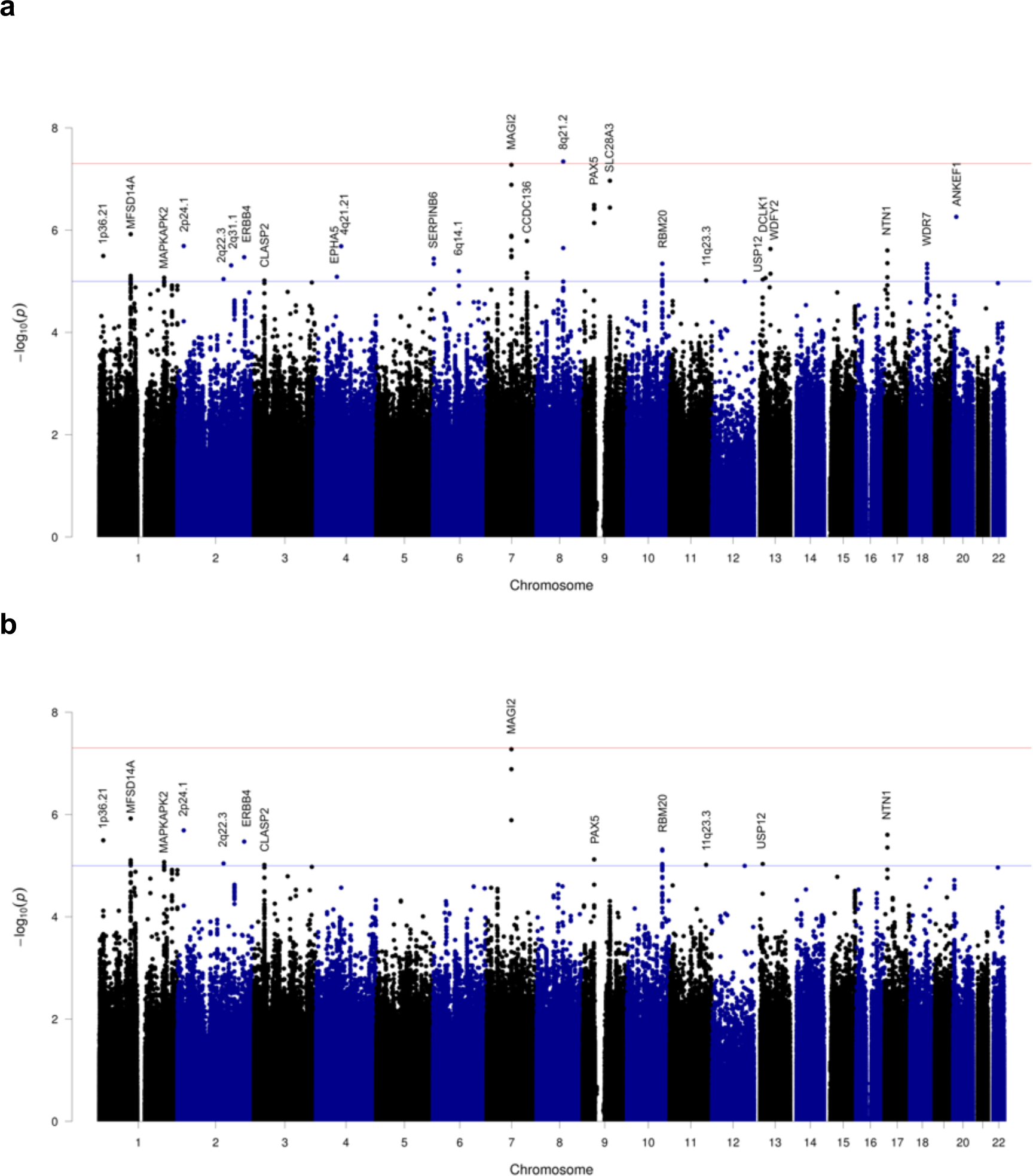
Manhattan plot for the genome-wide meta-analysis. Directly genotyped and imputed variants at MAF greater than 5% were tested for association with invasive GAS susceptibility in all four case-control analyses using linear mixed models. The association statistics were combined by **(a)** fixed-effects or **(b)** random-effects meta-analysis. Each point represents a single variant. The blue horizontal line indicates the suggestive significance threshold (*P* = 10^−5^) and the red horizontal line indicates the genome-wide significance threshold (*P* = 5 x 10^−8^).

**Suppl. Figure 7.**
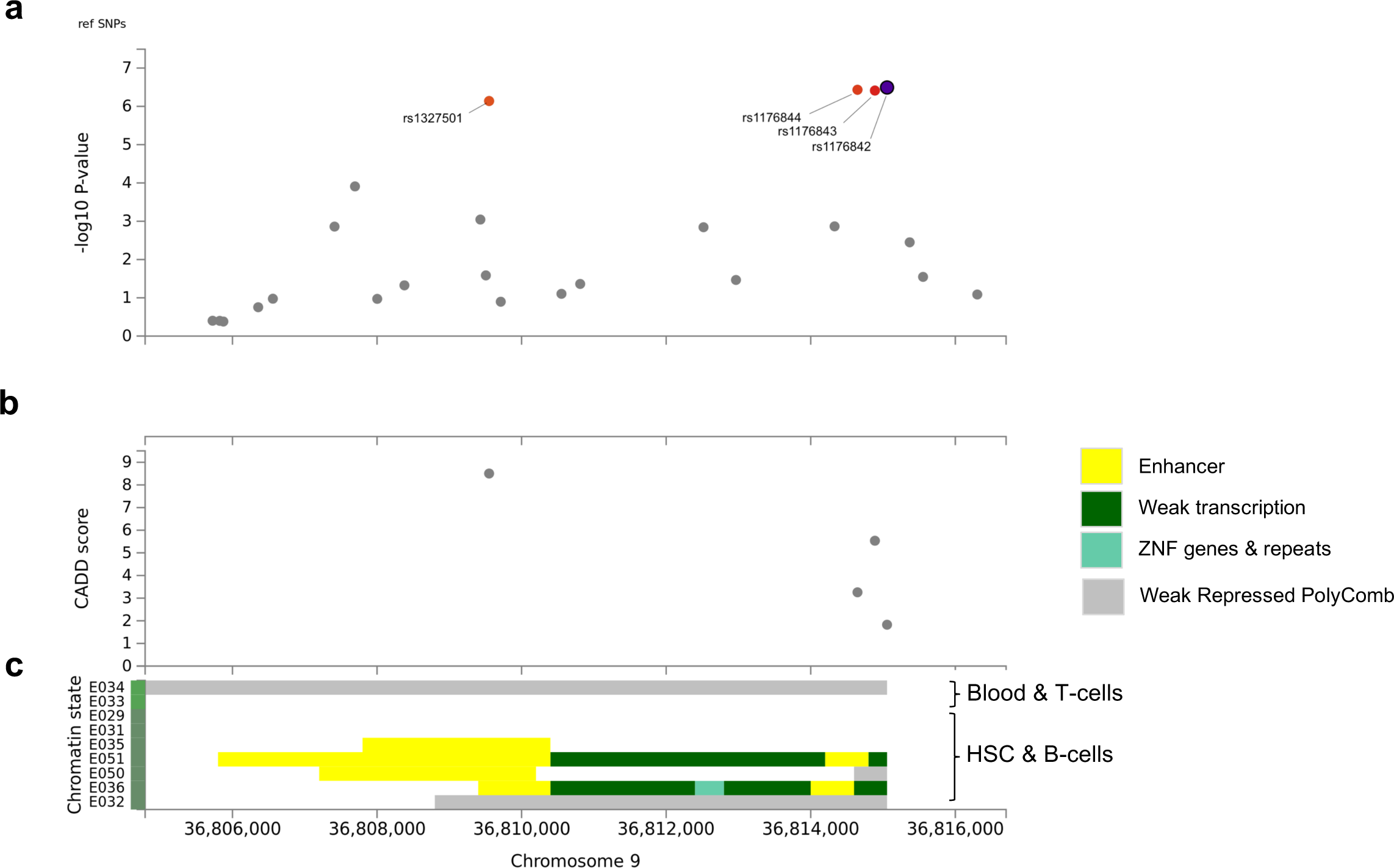
Putative regulatory effects of *PAX5* locus variants. Genomic position is plotted against: **(a)** the negative common logarithm of the p-value from the fixed-effects meta-analysis; **(b)** the Combined Annotation Dependent Depletion (CADD) score, a scaled measure of pathogenicity in which scores of 10 and 20, respectively, indicate that a variant is among the most 10% and 1% deleterious in the human genome; [32] **(c)** the chromatin states of nine cell types annotated using the core 15-state state model (ChromHMM), which predicts regulatory annotations based on histone modifications. [33] E034, primary T-cells from peripheral blood; E033, primary T cells from cord blood; E029, primary monocytes from peripheral blood; E031, primary B cells from cord blood; E035, primary hematopoietic stem cells; E051, primary hematopoietic stem cells G-CSF-mobilised (male); E050, primary hematopoietic stem cells G-CSF-mobilised (female); E036, primary hematopoietic stem cells in short term culture; E032, primary B cells from peripheral blood. The plot was generated using the FUMA web application.[34]

**Suppl. Table 1.**
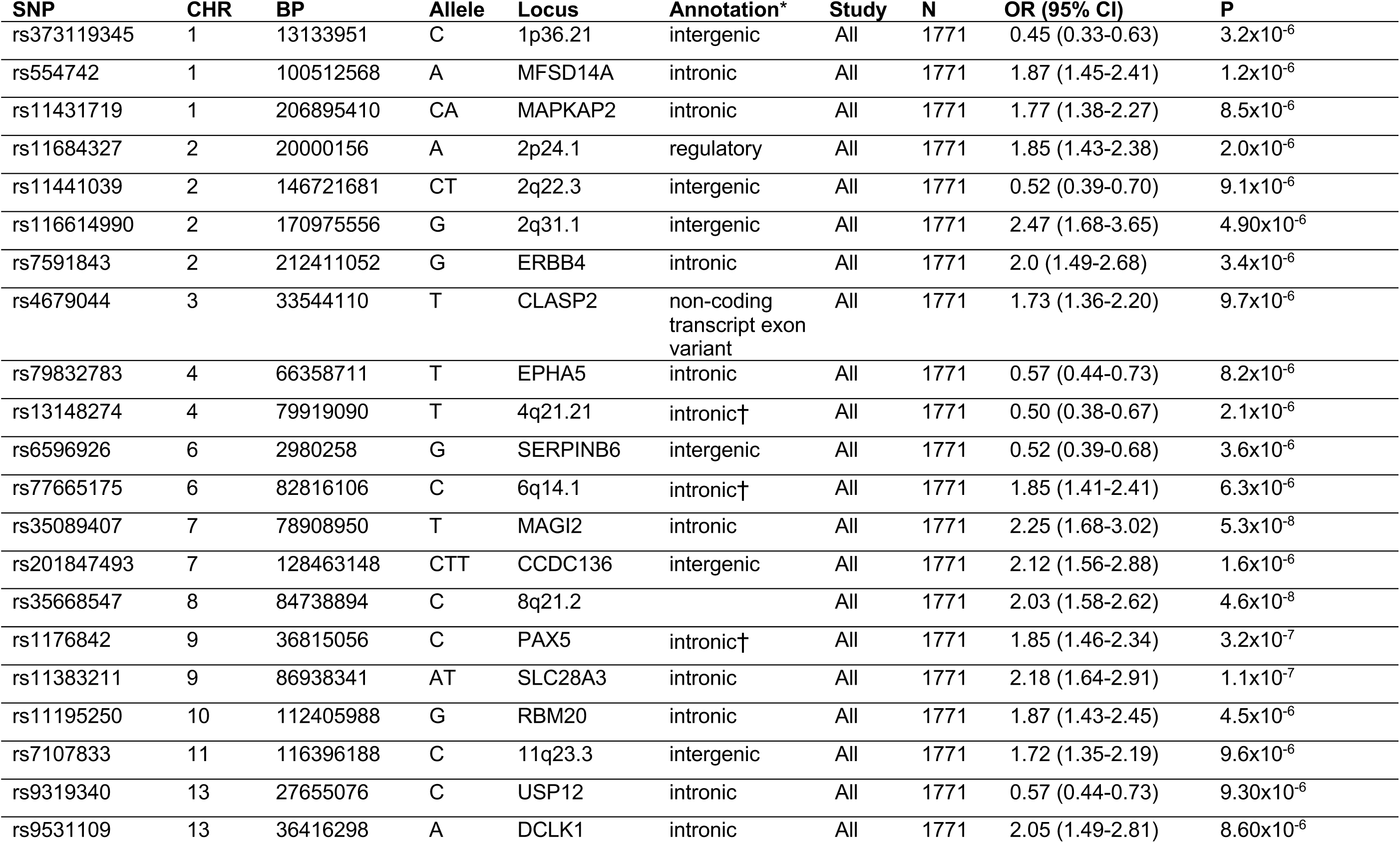
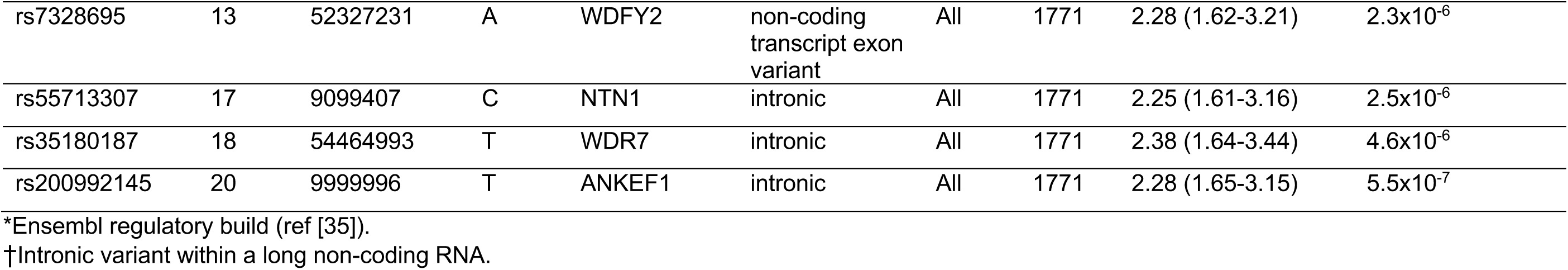
Genomic loci associated with susceptibility in a fixed-effects meta-analysis.

**Suppl. Table 2.** Genomic loci associated with susceptibility in a random-effects meta-analysis.

| SNP | CHR | BP | Allele | Locus | Annotation* | Study | N | MAF | Model | OR (95% CI) | P |
| --- | --- | --- | --- | --- | --- | --- | --- | --- | --- | --- | --- |
| rs373119345 | 1 | 13133951 | C | 1p36.21 | intergenic | Invasive (UK) | 473 | 0.72 | LMM | 0.38 (0.19-0.75) | 0.0054 |
|  |  |  |  |  |  | Emphy. (UK) | 396 | 0.74 | LMM | 0.37 (0.18-0.76) | 0.0071 |
|  |  |  |  |  |  | Bacte. (KE) | 726 | 0.63 | LMM | 0.57 (0.35-0.93) | 0.0254 |
|  |  |  |  |  |  | Invasive (NC) | 176 | 0.79 | LMM | 0.38 (0.13-1.11) | 0.082 |
|  |  |  |  |  |  | All | 1771 | - | FE | 0.45 (0.33-0.63) | 3.2x10<sup>-6</sup> |
|  |  |  |  |  |  |  |  | - | RE | 0.45 (0.33-0.63) | 3.2x10<sup>-6</sup> |
| rs554742 | 1 | 100512568 | A | MFSD14A | intronic | Invasive (UK) | 473 | 0.16 | LMM | 1.83 (1.08-3.08) | 0.0238 |
|  |  |  |  |  |  | Emphy. (UK) | 396 | 0.16 | LMM | 2.60 (1.54-4.38) | 0.0004 |
|  |  |  |  |  |  | Bacte. (KE) | 726 | 0.37 | LMM | 1.59 (1.10-2.29) | 0.0136 |
|  |  |  |  |  |  | Invasive (NC) | 176 | 0.09 | LMM | 2.04 (0.73-5.65) | 0.1813 |
|  |  |  |  |  |  | All | 1771 | - | FE | 1.87 (1.45-2.41) | 1.2x10<sup>-6</sup> |
|  |  |  |  |  |  |  |  | - | RE | 1.87 (1.45-2.41) | 1.2x10<sup>-6</sup> |
| rs11431719 | 1 | 206895410 | CA | MAPKAP2 | intronic | Invasive (UK) | 473 | 0.21 | LMM | 1.72 (1.04-2.83) | 0.0332 |
|  |  |  |  |  |  | Emphy. (UK) | 396 | 0.2 | LMM | 2.11 (1.21-3.70) | 0.0089 |
|  |  |  |  |  |  | Bacte. (KE) | 726 | 0.23 | LMM | 1.87 (1.28-2.74) | 0.0012 |
|  |  |  |  |  |  | Invasive (NC) | 176 | 0.48 | LMM | 1.08 (0.51-2.30) | 0.8485 |
|  |  |  |  |  |  | All | 1771 | - | FE | 1.77 (1.38-2.27) | 8.5x10<sup>-6</sup> |
|  |  |  |  |  |  |  |  | - | RE | 1.77 (1.38-2.27) | 8.5x10<sup>-6</sup> |
| rs11684327 | 2 | 20000156 | A | 2p24.1 | regulatory | Invasive (UK) | 473 | 0.18 | LMM | 2.24 (1.35-3.72) | 0.0017 |
|  |  |  |  |  |  | Emphy. (UK) | 396 | 0.18 | LMM | 1.63 (0.90-2.95) | 0.1082 |
|  |  |  |  |  |  | Bacte. (KE) | 726 | 0.27 | LMM | 1.59 (1.08-2.33) | 0.0189 |
|  |  |  |  |  |  | Invasive (NC) | 176 | 0.25 | LMM | 2.46 (1.24-4.88) | 0.0112 |
|  |  |  |  |  |  | All | 1771 | - | FE | 1.85 (1.43-2.38) | 2.0x10<sup>-6</sup> |
|  |  |  |  |  |  |  |  | - | RE | 1.85 (1.43-2.38) | 2.0x10<sup>-6</sup> |
| rs11441039 | 2 | 146721681 | CT | 2q22.3 | intergenic | Invasive (UK) | 473 | 0.59 | LMM | 0.54 (0.32-0.92) | 0.0233 |
|  |  |  |  |  |  | Empy. (UK) | 396 | 0.57 | LMM | 0.55 (0.31-0.98) | 0.0423 |
|  |  |  |  |  |  | Bacte. (KE) | 726 | 0.27 | LMM | 0.52 (0.31-0.87) | 0.0128 |
|  |  |  |  |  |  | Invasive (NC) | 176 | 0.52 | LMM | 0.44 (0.20-0.94) | 0.0374 |
|  |  |  |  |  |  | All | 1771 | - | FE | 0.52 (0.39-0.70) | 9.1x10<sup>-6</sup> |
|  |  |  |  |  |  |  |  | - | RE | 0.52 (0.39-0.70) | 9.1x10<sup>-6</sup> |
| rs7591843 | 2 | 212411052 | G | ERBB4 | intronic | Invasive (UK) | 473 | 0.19 | LMM | 2.28 (1.27-4.10) | 0.0056 |
|  |  |  |  |  |  | Empy. (UK) | 396 | 0.21 | LMM | 2.03 (1.12-3.69) | 0.0197 |
|  |  |  |  |  |  | Bacte. (KE) | 726 | 0.47 | LMM | 1.91 (1.26-2.89) | 0.0022 |
|  |  |  |  |  |  | Invasive (NC) | 176 | 0.06 | LMM | 0.30 (0.01-9.49) | 0.5053 |
|  |  |  |  |  |  | All | 1771 | - | FE | 2.0 (1.49-2.68) | 3.4x10<sup>-6</sup> |
|  |  |  |  |  |  |  |  | - | RE | 2.0 (1.49-2.68) | 3.4x10<sup>-6</sup> |
| rs4679044 | 3 | 33544110 | T | CLASP2 | non-coding<br>transcript<br>exon variant | Invasive (UK) | 473 | 0.37 | LMM | 2.13 (1.35-3.39) | 0.0012 |
|  |  |  |  |  |  | Empy. (UK) | 396 | 0.35 | LMM | 1.77 (1.09-2.88) | 0.0225 |
|  |  |  |  |  |  | Bacte. (KE) | 726 | 0.26 | LMM | 1.52 (1.03-2.24) | 0.0343 |
|  |  |  |  |  |  | Invasive (NC) | 176 | 0.64 | LMM | 1.47 (0.64-3.36) | 0.3684 |
|  |  |  |  |  |  | All | 1771 | - | FE | 1.73 (1.36-2.20) | 9.7x10<sup>-6</sup> |
|  |  |  |  |  |  |  |  | - | RE | 1.73 (1.36-2.20) | 9.7x10<sup>-6</sup> |
| rs35089407 | 7 | 78908950 | T | MAGI2 | intronic | Invasive (UK) | 473 | 0.09 | LMM | 1.46 (0.69-3.10) | 0.3248 |
|  |  |  |  |  |  | Empy. (UK) | 396 | 0.08 | LMM | 1.72 (0.77-3.83) | 0.1894 |
|  |  |  |  |  |  | Bacte. (KE) | 726 | 0.22 | LMM | 2.53 (1.74-3.69) | 1.3x10<sup>-6</sup> |
|  |  |  |  |  |  | Invasive (NC) | 176 | 0.1 | LMM | 2.95 (1.23-7.12) | 0.0179 |
|  |  |  |  |  |  | All | 1771 | - | FE | 2.25 (1.68-3.02) | 5.3x10<sup>-8</sup> |
|  |  |  |  |  |  |  |  | - | RE | 2.25 (1.68-3.02) | 5.3x10<sup>-8</sup> |
| rs1176844 | 9 | 36814647 | C | PAX5 | intronic† | Invasive (UK) | 473 | 0.48 | LMM | 2.69 (1.67-4.35) | 4.9x10<sup>-5</sup> |
|  |  |  |  |  |  | Empy. (UK) | 396 | 0.47 | LMM | 1.67 (1.04-2.68) | 0.0346 |
|  |  |  |  |  |  | Bacte. (KE) | 726 | 0.41 | LMM | 1.50 (1.04-2.17) | 0.0305 |
|  |  |  |  |  |  | Invasive (NC) | 176 | 0.33 | LMM | 2.07 (1.03-4.18) | 0.0457 |
|  |  |  |  |  |  | All | 1771 | - | FE | 1.84 (1.45-2.32) | 3.7x10<sup>-7</sup> |
|  |  |  |  |  |  |  |  | - | RE | 1.86 (1.42-2.44) | 7.6x10<sup>-6</sup> |
| rs11195250 | 10 | 112405988 | G | RBM20 | intronic | Invasive (UK) | 473 | 0.09 | LMM | 2.54 (1.39-4.63) | 0.0024 |
|  |  |  |  |  |  | Empy. (UK) | 396 | 0.11 | LMM | 2.36 (1.25-4.46) | 0.0081 |
|  |  |  |  |  |  | Bacte. (KE) | 726 | 0.39 | LMM | 1.50 (1.04-2.17) | 0.0314 |
|  |  |  |  |  |  | Invasive (NC) | 176 | 0.15 | LMM | 2.21 (0.93-5.29) | 0.0793 |
|  |  |  |  |  |  | All | 1771 | - | FE | 1.87 (1.43-2.45) | 4.5x10<sup>-6</sup> |
|  |  |  |  |  |  |  |  | - | RE | 1.88 (1.43-2.46) | 4.8x10<sup>-6</sup> |
| rs7107833 | 11 | 116396188 | C | 11q23.3 | intergenic | Invasive (UK) | 473 | 0.32 | LMM | 2.03 (1.29-3.20) | 0.0022 |
|  |  |  |  |  |  | Empy. (UK) | 396 | 0.31 | LMM | 1.64 (0.96-2.80) | 0.0705 |
|  |  |  |  |  |  | Bacte. (KE) | 726 | 0.29 | LMM | 1.48 (1.02-2.14) | 0.0411 |
|  |  |  |  |  |  | Invasive (NC) | 176 | 0.28 | LMM | 2.30 (1.07-4.94) | 0.0369 |
|  |  |  |  |  |  | All | 1771 | - | FE | 1.72 (1.35-2.19) | 9.6x10<sup>-6</sup> |
|  |  |  |  |  |  |  |  | - | RE | 1.72 (1.35-2.19) | 9.6x10<sup>-6</sup> |
| rs9319340 | 13 | 27655076 | C | USP12 | intronic | Invasive (UK) | 473 | 0.83 | LMM | 0.61 (0.35-1.08) | 0.0866 |
|  |  |  |  |  |  | Empy. (UK) | 396 | 0.8 | LMM | 0.44 (0.27-0.71) | 0.0009 |
|  |  |  |  |  |  | Bacte. (KE) | 726 | 0.41 | LMM | 0.66 (0.44-0.97) | 0.0346 |
|  |  |  |  |  |  | Invasive (NC) | 176 | 0.56 | LMM | 0.53 (0.26-1.10) | 0.0948 |
|  |  |  |  |  |  | All | 1771 | - | FE | 0.57 (0.44-0.73) | 9.30x10<sup>-6</sup> |
|  |  |  |  |  |  |  |  | - | RE | 0.57 (0.44-0.73) | 9.30x10<sup>-6</sup> |
| rs55713307 | 17 | 9099407 | C | NTN1 | intronic | Invasive (UK) | 473 | 0.11 | LMM | 3.22 (1.62-6.39) | 0.0008 |
|  |  |  |  |  |  | Emphy. (UK) | 396 | 0.1 | LMM | 2.34 (1.09-5.02) | 0.0303 |
|  |  |  |  |  |  | Bacte. (KE) | 726 | 0.13 | LMM | 1.83 (1.11-3.04) | 0.0187 |
|  |  |  |  |  |  | Invasive (NC) | 176 | 0.16 | LMM | 2.22 (0.81-6.13) | 0.1294 |
|  |  |  |  |  |  | All | 1771 | - | FE | 2.25 (1.61-3.16) | 2.5x10<sup>-6</sup> |
|  |  |  |  |  |  |  |  | - | RE | 2.25 (1.61-3.16) | 2.5x10<sup>-6</sup> |

\*Ensembl regulatory build (ref [35]).
†Intronic variant within a long non-coding RNA.
KE, Kenya; NC, New Caledonia; MAF, minor allele frequency; LMM, linear mixed model; FE, fixed-effects meta-analysis; RE, random-effects meta-analysis.

**Suppl. Table 3.** Sensitivity analysis to identify the most robust association signals.

| SNP | CHR | BP | Allele | Locus | Initial P<sub>FE</sub> | Initial P<sub>RE</sub> | Min P<sub>FE</sub> | Max P<sub>FE</sub> | RegulomeDB score* |
| --- | --- | --- | --- | --- | --- | --- | --- | --- | --- |
| rs11441039 | 2 | 146721681 | CT | 2q22.3 | 9.1x10<sup>-6</sup> | 9.1x10<sup>-6</sup> | 8.2x10<sup>-7</sup> | 7.4x10<sup>-5</sup> | 6 |
| rs7591843 | 2 | 212411052 | G | ERBB4 | 3.4x10<sup>-6</sup> | 3.4x10<sup>-6</sup> | 1.9x10<sup>-7</sup> | 7.0 x10<sup>-5</sup> | 6 |
| rs4679044 | 3 | 33544110 | T | CLASP2 | 9.7x10<sup>-6</sup> | 9.7x10<sup>-6</sup> | 4.4x10<sup>-7</sup> | 8.8x10<sup>-5</sup> | 6 |
| rs62460858 | 7 | 78900116 | C | MAGI2 | 1.3x10<sup>-7</sup> | 1.3x10<sup>-8</sup> | 1.5x10<sup>-8</sup> | 6.0x10<sup>-5</sup> | No data |
| rs35089407 | 7 | 78908950 | T | MAGI2 | 5.3x10<sup>-8</sup> | 5.3x10<sup>-8</sup> | 1.2x10<sup>-8</sup> | 3.1x10<sup>-5</sup> | No data |
| rs1327501 | 9 | 36809551 | A | PAX5 | 7.3x10<sup>-7</sup> | 2.4x10<sup>-5</sup> | 1.9x10<sup>-7</sup> | 4.5x10<sup>-5</sup> | 4 |
| rs1176844 | 9 | 36814647 | C | PAX5 | 3.7x10<sup>-7</sup> | 7.6x10<sup>-6</sup> | 1.3x10<sup>-7</sup> | 3.5x10<sup>-5</sup> | 6 |
| rs1176843 | 9 | 36814888 | C | PAX5 | 3.9x10<sup>-7</sup> | 6.7x10<sup>-5</sup> | 1.1x10<sup>-7</sup> | 3.0x10<sup>-5</sup> | 6 |
| rs1176842 | 9 | 36815056 | C | PAX5 | 3.2x10<sup>-7</sup> | 6.0x10<sup>-5</sup> | 1.2x10<sup>-7</sup> | 3.3x10<sup>-5</sup> | 5 |

\*Range 1–6 with 4 indicating “Transcription factor binding and DNase hypersensitivity peak”, 5 indicating “Transcription factor binding or DNase hypersensitivity peak”, and 6 indicating “Other evidence” (see: http://www.regulomedb.org/help).

## Notes

### Competing Interest Statement

The authors have declared no competing interest.

### Author Declarations

All relevant ethical guidelines have been followed and any necessary IRB and/or ethics committee approvals have been obtained.

Any clinical trials involved have been registered with an ICMJE-approved registry such as ClinicalTrials.gov and the trial ID is included in the manuscript.

## References

1. Lamagni TL, Darenberg J, Luca-Harari B, et al. Epidemiology of severe Streptococcus pyogenes disease in Europe. J Clin Microbiol 2008; 46:2359–2367.

2. Carapetis JR, Steer AC, Mulholland EK, Weber M. The global burden of group A streptococcal diseases. Lancet Infect Dis 2005; 5:685–694. Available at: http://eutils.ncbi.nlm.nih.gov/entrez/eutils/elink.fcgi?dbfrom=pubmed&id=16253886&retmode=ref&cmd=prlinks.

3. Musser JM, Shelburne SA. A decade of molecular pathogenomic analysis of group A Streptococcus. J Clin Invest 2009; 119:2455–2463.

4. Steer AC, Lamagni T, Curtis N, Carapetis JR. Invasive group a streptococcal disease: epidemiology, pathogenesis and management. Drugs 2012; 72:1213–1227. Available at: http://www.ncbi.nlm.nih.gov/pubmed/22686614.

5. Engel ME, Stander R, Vogel J, Adeyemo AA, Mayosi BM. Genetic Susceptibility to Acute Rheumatic Fever: A Systematic Review and Meta-Analysis of Twin Studies. PLoS ONE 2011; 6:e25326.

6. Kvestad E, Kvaerner KJ, Røysamb E, Tambs K, Harris JR, Magnus P. Heritability of recurrent tonsillitis. Arch. Otolaryngol. Head Neck Surg. 2005; 131:383–387.

7. Kotb M, Norrby-Teglund A, McGeer A, et al. An immunogenetic and molecular basis for differences in outcomes of invasive group A streptococcal infections. Nat Med 2002; 8:1398–1404.

8. Parks T, Elliott K, Lamagni T, et al. Elevated risk of invasive group A streptococcal disease and host genetic variation in the human leukocyte antigen locus. Genes Immun 2019;

9. Imanishi K, Igarashi H, Uchiyama T. Relative abilities of distinct isotypes of human major histocompatibility complex class II molecules to bind streptococcal pyrogenic exotoxin types A and B. Infect Immun 1992; 60:5025–5029.

10. Llewelyn M, Sriskandan S, Peakman M, et al. HLA class II polymorphisms determine responses to bacterial superantigens. J Immunol 2004; 172:1719–1726.

11. Davila S, Wright VJ, Khor CC, et al. Genome-wide association study identifies variants in the CFH region associated with host susceptibility to meningococcal disease. Nat Genet 2010; 42:772–776.

12. Kenyan Bacteraemia Study Group, Wellcome Trust Case Control Consortium 2 (WTCCC2), Rautanen A, et al. Polymorphism in a lincRNA Associates with a Doubled Risk of Pneumococcal Bacteremia in Kenyan Children. Am J Hum Genet 2016; 98:1092–1100.

13. Gilchrist JJ, Rautanen A, Fairfax BP, et al. Risk of nontyphoidal Salmonella bacteraemia in African children is modified by STAT4. Nat Commun 2018; 9:2489.

14. Parks T, Elliott K, Lamagni T, et al. Elevated risk of invasive group A streptococcal disease and host genetic variation in the human leukocyte antigen locus. 2019; Available at: http://dx.doi.org/10.1101/559161.

15. Berkley JA, Lowe BS, Mwangi I, et al. Bacteremia among children admitted to a rural hospital in Kenya. N Engl J Med 2005; 352:39–47.

16. Parks T, Mirabel MM, Kado J, et al. Association between a common immunoglobulin heavy chain allele and rheumatic heart disease risk in Oceania. Nat Commun 2017; 8:14946.

17. Anderson CA, Pettersson FH, Clarke GM, Cardon LR, Morris AP, Zondervan KT. Data quality control in genetic case-control association studies. Nat Protoc 2010; 5:1564–1573.

18. Lee SH, Nyholt DR, Macgregor S, et al. A Simple and Fast Two-Locus Quality Control Test to Detect False Positives Due to Batch Effects in Genome-Wide Association Studies. Genet Epidemiol 2010; 34:854–862.

19. Yang J, Lee SH, Goddard ME, Visscher PM. GCTA: A tool for genome-wide complex trait analysis. Am J Hum Genet 2011; 88:76–82.

20. Han B, Eskin E. Random-Effects Model Aimed at Discovering Associations in Meta-Analysis of Genome-wide Association Studies. Am J Hum Genet 2011; 88:586–598.

21. Lloyd-Jones LR, Robinson MR, Yang J, Visscher PM. Transformation of Summary Statistics from Linear Mixed Model Association on All-or-None Traits to Odds Ratio. Genetics 2018; 208:1397–1408.

22. Boyle AP, Hong EL, Hariharan M, et al. Annotation of functional variation in personal genomes using RegulomeDB. Genome Res 2012; 22:1790–1797.

23. Cobaleda C, Schebesta A, Delogu A, Busslinger M. Pax5: the guardian of B cell identity and function. Nat Immunol 2007; 8:463–470.

24. Norrby-Teglund A, Pauksens K, Holm SE, Norgren M. Relation between low capacity of human sera to inhibit streptococcal mitogens and serious manifestation of disease. J Infect Dis 1994; 170:585–591.

25. Norrby-Teglund A, Kaul R, Low DE, et al. Evidence for the presence of streptococcal-superantigen-neutralizing antibodies in normal polyspecific immunoglobulin G. Infect Immun 1996; 64:5395–5398.

26. Parks T, Wilson C, Curtis N, Norrby-Teglund A, Sriskandan S. Polyspecific intravenous immunoglobulin in clindamycin-treated patients with streptococcal toxic shock syndrome: a systematic review and meta-analysis. Clin Infect Dis 2018; 1:69–1436.

27. Bauß K, Knapp B, Jores P, et al. Phosphorylation of the Usher syndrome 1G protein SANS controls Magi2-mediated endocytosis. Hum Mol Genet 2014; 23:3923–3942.

28. Marshall CR, Young EJ, Pani AM, et al. Infantile spasms is associated with deletion of the MAGI2 gene on chromosome 7q11.23-q21.11. Am J Hum Genet 2008; 83:106–111.

29. Kathiresan S, Manning AK, Demissie S, et al. A genome-wide association study for blood lipid phenotypes in the Framingham Heart Study. BMC Med Genet 2007; 8 Suppl 1:S17.

30. Li YR, Keating BJ. Trans-ethnic genome-wide association studies: advantages and challenges of mapping in diverse populations. Genome Med 2014; 6:91.

31. Marchini J, Howie B. Genotype imputation for genome-wide association studies. Nat Rev Genet 2010; 11:499–511. Available at: http://www.nature.com/nrg/journal/v11/n7/full/nrg2796.html.

32. Kircher M, Witten DM, Jain P, O’Roak BJ, Cooper GM, Shendure J. A general framework for estimating the relative pathogenicity of human genetic variants. Nat Genet 2014; 46:310–315.

33. Roadmap Epigenomics Consortium, Kundaje A, Meuleman W, et al. Integrative analysis of 111 reference human epigenomes. Nature 2015; 518:317–330.

34. Watanabe K, Taskesen E, van Bochoven A, Posthuma D. Functional mapping and annotation of genetic associations with FUMA. Nat Commun 2017; 8:1826.

35. Zerbino DR, Wilder SP, Johnson N, Juettemann T, Flicek PR. The ensembl regulatory build. Genome Biol 2015; 16:56.

